# Estimated prevalence of dental fear in adults: a systematic review and meta-analysis

**DOI:** 10.1101/2020.11.30.20241216

**Authors:** Ethieli R. Silveira, Mariana G. Cademartori, Helena S. Schuch, Jason M. Armfield, Flavio F. Demarco

## Abstract

**Objectives:** To systematically review the literature on the prevalence of dental fear in adults to estimate a worldwide pooled prevalence and to investigate factors related to the heterogeneity of estimates.

**Methods:** Inclusion criteria were population-based studies reporting the prevalence or data that allowed the calculation of prevalence of dental fear in adults and/or elders. Five electronic databases (Embase, PubMed, Scopus, Virtual Health Library (BVS) and Web of Science) were searched. Two researchers independently performed the study selection, data extraction and quality assessment of the included studies. The prevalence pooled estimates of dental fear were calculated using fixed- and random-effect models. Subgroup analyses were performed to investigate variability between studies.

**Results:** The search strategy identified 4,486 studies. After removal of duplicates (1,722), title and abstract screening (2,764) and full-text reading (108), 31 publications were deemed eligible for this systematic review. A total of 72,577 individuals 18 years of age or older composed the sample of this systematic review. The global estimated prevalence of dental fear and anxiety (DFA), high DFA and severe DFA in adults were 13.8%, 11.2% and 2.6%, respectively. Subgroup analyses showed a higher prevalence of DFA, high DFA and severe DFA among women and younger adults. The instrument used to measure dental fear also affected the prevalence of the outcome.

**Conclusions:** Dental fear and high dental fear are prevalent in adults worldwide, being more prevalent among women.

## INTRODUCTION

Previous studies have proposed and demonstrated the theory of a vicious circle of dental fear^1^. This theory postulates that fearful individuals tend to avoid dental visits, which may lead to impaired oral health and dental pain. This lack of routine or preventive dental check-ups means that, when they do visit the dentist, more invasive treatments are usually required, consequently increasing their dental fear^1, 2^.

The development of dental fear can originate in childhood, adolescence or even later in life^3^. The prevalence of dental fear among adults varies widely in the literature, with reports ranging from 4.2%^4^ to over than 50%^5^. These differences may reflect the great cultural, social and economic differences between populations, as well as the characteristics of the study, such as design and instrument used to measure dental fear^6, 7^. Numerous factors may be related to the presence of dental fear, ranging from individual characteristics such as age, gender and previous dental experience; to environment and socioeconomic conditions^8-10^. Also, there are several instruments to evaluate the presence of dental fear, varying from a single question to more complex scales, which could contribute to differences in prevalence^6^.

Systematic reviews are a useful tool to gather information from different studies to aggregate these data and provide evidence to researchers and clinicians. A single meta-analysis has previously investigated the global prevalence of dental fear in children and adolescents^10^. The authors observed that the prevalence of dental fear/anxiety varied considerably (10.0-29.3%) depending on the scale used to determine the outcome^11^. Global estimates for a determined outcome are useful for dental health systems in order to estimate the true magnitude of the problem and to establish preventive measurement or to provide treatment^12^. Until now, there have been no systematic reviews evaluating dental fear occurrence in adults.

Figures on oral conditions observed worldwide are alarming, with challenging situations observed for adults and especially older adults^13^. Since dental fear is considered a barrier for seeking dental treatment, which may lead to worsening the oral condition, investigating the prevalence of dental fear in adults, including older adults, is important. Thus, the aim of this study was to systematically review the literature on the prevalence of dental fear in adults, and older adults specifically, to estimate a worldwide pooled prevalence and to investigate factors related to the heterogeneity of estimates.

## MATERIAL AND METHODS

This study was reported according to the Preferred Reporting Items for Systematic Reviews and Meta-Analyses (PRISMA) guidelines^14^.

### Review question

The review question was defined according to the PICO strategy: (a) P: representative population-based studies; (b) I: adults (≥18 years) with dental fear; (c) C: adults (≥18 years) without dental fear; (d) O: dental fear prevalence. Thus, the following review question formed the basis of this systematic review: ‘What is the global estimated prevalence of dental fear in adults?’.

### Eligibility criteria

Original observational studies reporting the prevalence or data that allowed the calculation of prevalence of dental fear in adults were considered for inclusion. To ensure representativeness of the sample, only population-based studies with representative samples were considered. Studies with specific samples, such as patients of a dental clinic or individuals with a specific health condition were excluded. In addition, observational studies with convenience samples or those whose prevalence data could not be obtained were excluded. Additionally, eligible studies required participants to be aged 18 years or older. Studies that comprised individuals younger than 18 years of age were only included if they presented stratified prevalence estimates or provided data for the calculation of this estimate excluding those participants under 18 years of age.

In relation to the evaluation of dental fear, eligible methods included self-report, assessment using validated tools or single-item questions, or even diagnosed in terms of the Diagnostic and Statistical Manual of Mental Disorders V (DSM-V) criteria for specific dental phobia^15^. Dental fear was defined according to three binary categorizations: ‘Any dental fear’, ‘High dental fear’, and ‘Extreme/severe dental fear and dental phobia’. Any dental fear was measured as any degree of fear, high dental fear included those with moderate to severe, and extreme dental fear was restricted to those with the most severe category of the problem. Taking the single-item Dental Anxiety Question (DAQ) as an example, for the “Any dental fear” evaluation, those reporting from a little to very afraid were considered as cases with ‘any dental fear’, while those reporting quite to very afraid were considered cases of ‘high dental fear’ and those reporting to be very afraid were considered to have ‘extreme dental fear’.

To calculate a global prevalence, some methodological decisions were required. Prevalence rates of dental fear were collected or calculated, if necessary. When estimates from two or more dental fear instruments were presented in the same manuscript, priority was given to the more widely used instruments in the scientific literature, starting from the validated scales and proceeding to single-item questions. In addition, when more than one category of dental fear estimate was presented, the most severe was chosen. When two or more publications were published with the same data (sample), priority was given to the most recently published with data on dental fear relative or absolute frequencies.

### Search strategy and data collection

An electronic search was conducted in Embase, PubMed, Scopus, Virtual Health Library (BVS) and Web of Science databases without language restrictions to identify articles published up to March 2020. An initial search was conducted on PubMed with a search strategy using relevant MeSH and free terms related to study design and dental fear. Search strategies for each database are presented in Table 1.

**Table 1.** Search strategies used according to specific databases. 2020

| Database | Search strategies | Hits |
| --- | --- | --- |
| Embase | ('cross-sectional stud*' OR prevalence* OR 'cohort stud*' OR 'incidence stud*' OR 'epidemiologic stud*' OR 'mass screening'/exp OR 'health survey*' OR 'longitudinal stud*' OR 'longitudinal survey*') AND ('dental anxiet*' OR 'dental fear*' OR 'dental phobia*' OR odontophobia*) | 641 |
| Virtual Health Library (BVS) | (tw:(("Cross-Sectional Studies" OR "Cross-Sectional Study" OR "Cross Sectional Studies" OR "Cross Sectional Study" OR "Prevalence Study" OR "Prevalence Studies" OR Prevalence OR Prevalences OR "Cohort Studies" OR "Cohort Study" OR "Incidence Studies" OR "Incidence Study" OR "Epidemiologic Studies" OR "Epidemiological Studies" OR "Epidemiological Study" OR "Epidemiologic Study" OR "Mass screening" OR "Dental Health Survey" OR "Dental Health Surveys" OR "Longitudinal Studies" OR "Longitudinal Study" OR "Longitudinal Surveys") )) AND (tw:(("Dental anxiety" OR "Dental fear" OR "Dental fears" OR Odontophobia OR "Dental Phobia" OR "Dental Phobias" OR "Dental Anxieties")))) | 827 |
| Scopus | TITLE-ABS-KEY ("Cross-Sectional Studies" OR "Cross-Sectional Study" OR "Cross Sectional Studies" OR "Cross Sectional Study" OR "Prevalence Study" OR "Prevalence Studies" OR Prevalence OR Prevalences OR "Cohort Studies" OR "Cohort Study" OR "Incidence Studies" OR "Incidence Study" OR "Epidemiologic Studies" OR "Epidemiological Studies" OR "Epidemiological Study" OR "Epidemiologic Study" OR "Mass screening" OR "Dental Health Survey" OR "Dental Health Surveys" OR "Longitudinal Studies" OR "Longitudinal Study" OR "Longitudinal Surveys") AND TITLE-ABS-KEY("Dental anxiety" OR "Dental fear" OR "Dental fears" OR Odontophobia OR "Dental Phobia" OR "Dental Phobias" OR "Dental Anxieties") | 656 |
| PubMed | ((((((((((((((((((((((((((((((((((("Cross-Sectional Studies"[MeSH Terms]) OR "Cross-Sectional Studies") OR "Cross-Sectional Study") OR "Cross Sectional Studies") OR "Cross Sectional Study") OR "Prevalence Study") OR "Prevalence Studies") OR Prevalence[MeSH Terms]) OR Prevalence) OR Prevalences) OR "Cohort Studies"[MeSH Terms]) OR "Cohort Studies") OR "Cohort Study") OR "Incidence Studies") OR "Incidence Study") OR "Epidemiologic Studies"[MeSH Terms]) OR "Epidemiologic Studies") OR "Epidemiological | 1,783 |
|  | Studies") OR "Epidemiological Study") OR "Epidemiologic Study") OR "Mass screening") OR "Mass screening"[MeSH Terms]) OR "Health Surveys"[MeSH Terms]) OR "Health Surveys") OR "Health Survey") OR "Dental Health Survey") OR "Dental Health Surveys"[MeSH Terms]) OR "Dental Health Surveys") OR "Longitudinal Studies"[MeSH Terms]) OR "Longitudinal Studies") OR "Longitudinal Study") OR "Longitudinal Survey") OR "Longitudinal Surveys")) AND (((((((("Dental anxiety"[MeSH Terms]) OR "Dental anxiety") OR "Dental fear") OR "Dental fears") OR Odontophobia) OR Odontophobia) OR "Dental Phobia") OR "Dental Phobias") OR "Dental Anxieties") |  |
| Web of Science | TS=("Cross-Sectional Studies" OR "Cross-Sectional Study" OR "Cross Sectional Studies" OR "Cross Sectional Study" OR "Prevalence Study" OR "Prevalence Studies" OR Prevalence OR Prevalences OR "Cohort Studies" OR "Cohort Study" OR "Incidence Studies" OR "Incidence Study" OR "Epidemiologic Studies" OR "Epidemiological Studies" OR "Epidemiological Study" OR "Epidemiologic Study" OR "Mass screening" OR "Dental Health Survey" OR "Dental Health Surveys" OR "Longitudinal Studies" OR "Longitudinal Study" OR "Longitudinal Surveys") AND TS=("Dental anxiety" OR "Dental fear" OR "Dental fears" OR Odontophobia OR "Dental Phobia" OR "Dental Phobias" OR "Dental Anxieties") | 579 |

Duplicate references were identified and subsequently excluded. Two reviewers (HSS and MGC) independently screened titles and abstracts. Lists of the selected studies were compared and, in case of disagreement, a third reviewer was consulted, and a consensus was reached by discussion. The same two reviewers independently read full texts of the articles considering the eligibility criteria pre-established. Excluded articles and reasons for the exclusion are described in Table 2. In the next step, the reviewers performed a hand search in the reference list of the included studies. Grey literature was searched using the Google Scholar search considering the first 100 hits. As in the first screening, in case of disagreements, a third reviewer was consulted, and a consensus was reached by discussion.

**Table 2.**
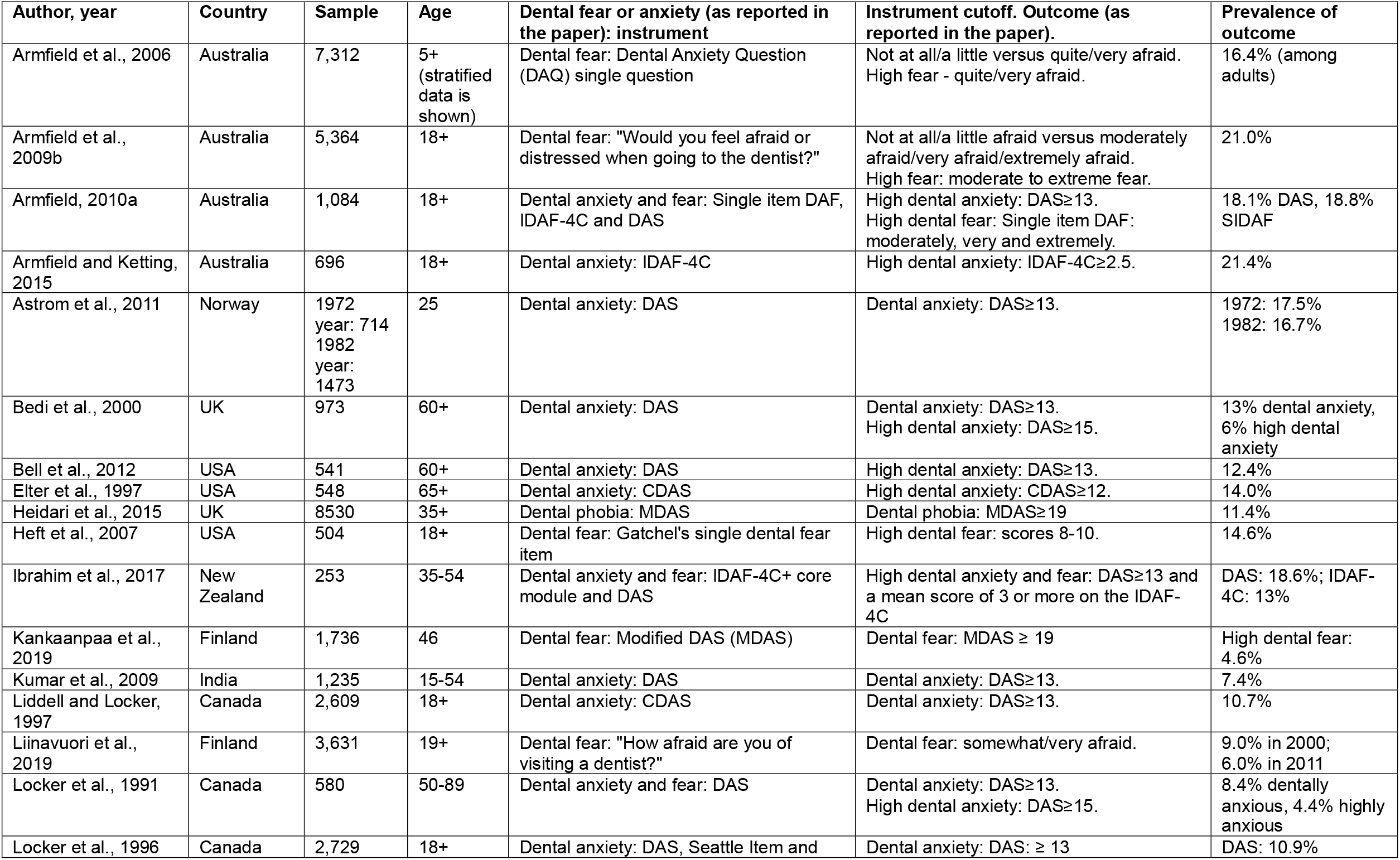
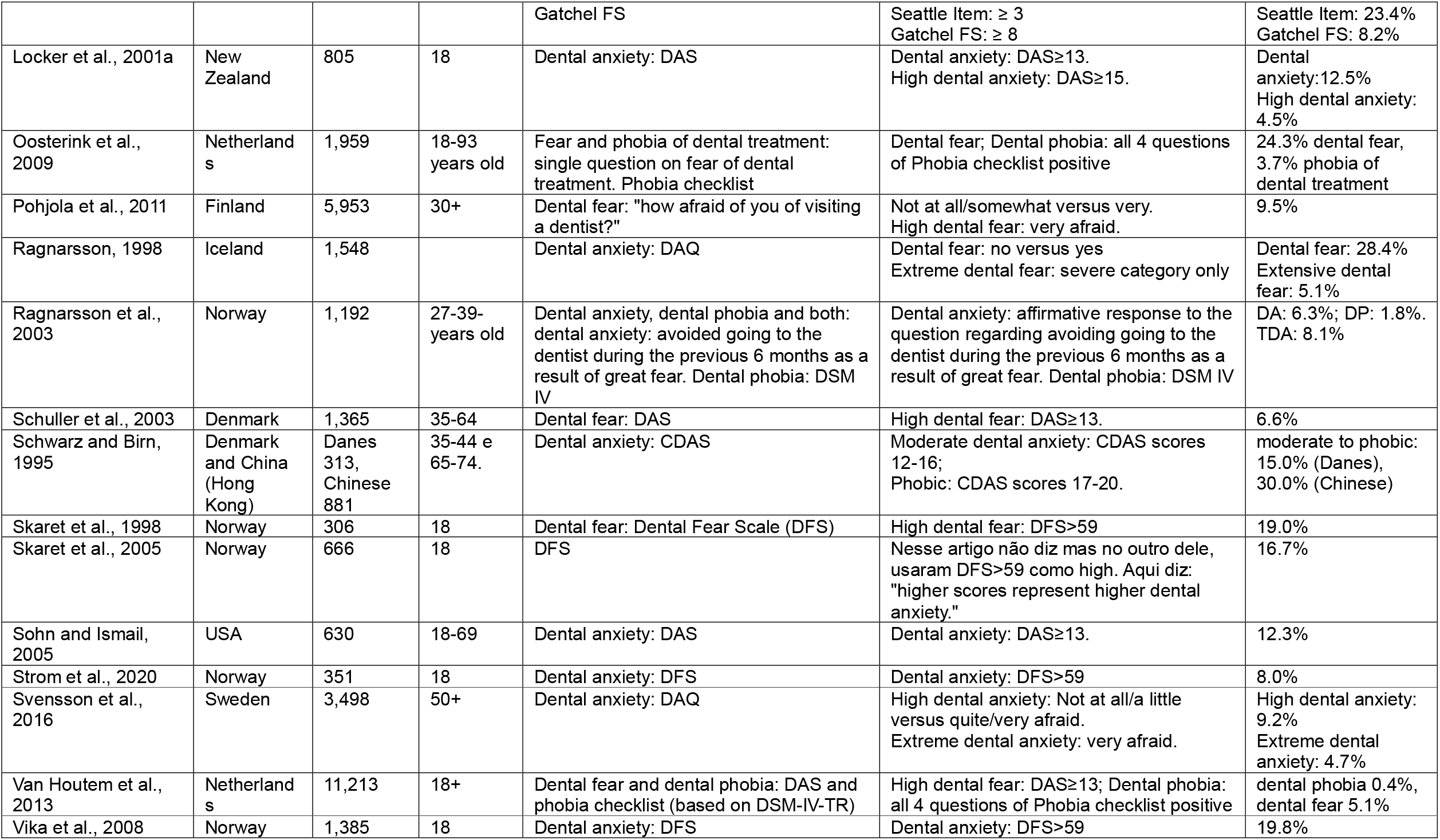
Main characteristics of included studies. 2020.

Data collection was independently performed by the same two reviewers (HSS and MGC) using a predefined worksheet. Characteristics of participants (gender and age), study design, sample size, instrument used to assess outcome, main results and conclusion were collected. In case of disagreement, a third reviewer was consulted, and a consensus was reached by discussion. When necessary, authors where contacted, and questions about the study were clarified.

### Critical appraisal

The methodological quality assessment of the included studies was performed using the Critical Appraisal Checklist for Prevalence and Incidence studies proposed by The Joanna Briggs Institute^16^. This instrument is comprised of 10 items which should be answered with ‘yes’, ‘no’, or ‘unclear’. The overall score for each study was identified through the sum of yes answers. Studies with scores from 0 to 5 were rated as high risk of bias, and those with scores ranging from 6 to 10 as low risk of bias. The same two reviewers conducted quality assessment, and disagreements were resolved by reaching consensus through discussion.

### Statistical analysis

The prevalence pooled estimates of dental fear were calculated using fixed- and random-effect models. The random-effects model was chosen in the presence of heterogeneity (I^2^>50%)^17^. To investigate if methodological characteristics influenced variability between studies, estimated prevalence was explored using subgroup analyses for each level of dental fear according to the assessment method, gender and age (younger and older adults) groups. Sensitivity analyses were conducted to observe the effect of each study on the pooled prevalence estimates. Data were analyzed using the software Stata 14.0 (Stata Corp, College Station, TX, USA).

## RESULTS

The search strategy identified 4,486 studies. From those, 1,722 were duplicates and therefore excluded. A total of 2,764 articles were screened for titles and abstracts. In the second screening, 108 studies received full-text reading, and 31 publications were deemed eligible for this systematic review. Main reasons for study exclusion are presented in Supplementary Table 1. Figure 1 presents the flowchart of study selection as recommended by the PRISMA Statement. Main characteristics of selected studies are presented in Table 2.

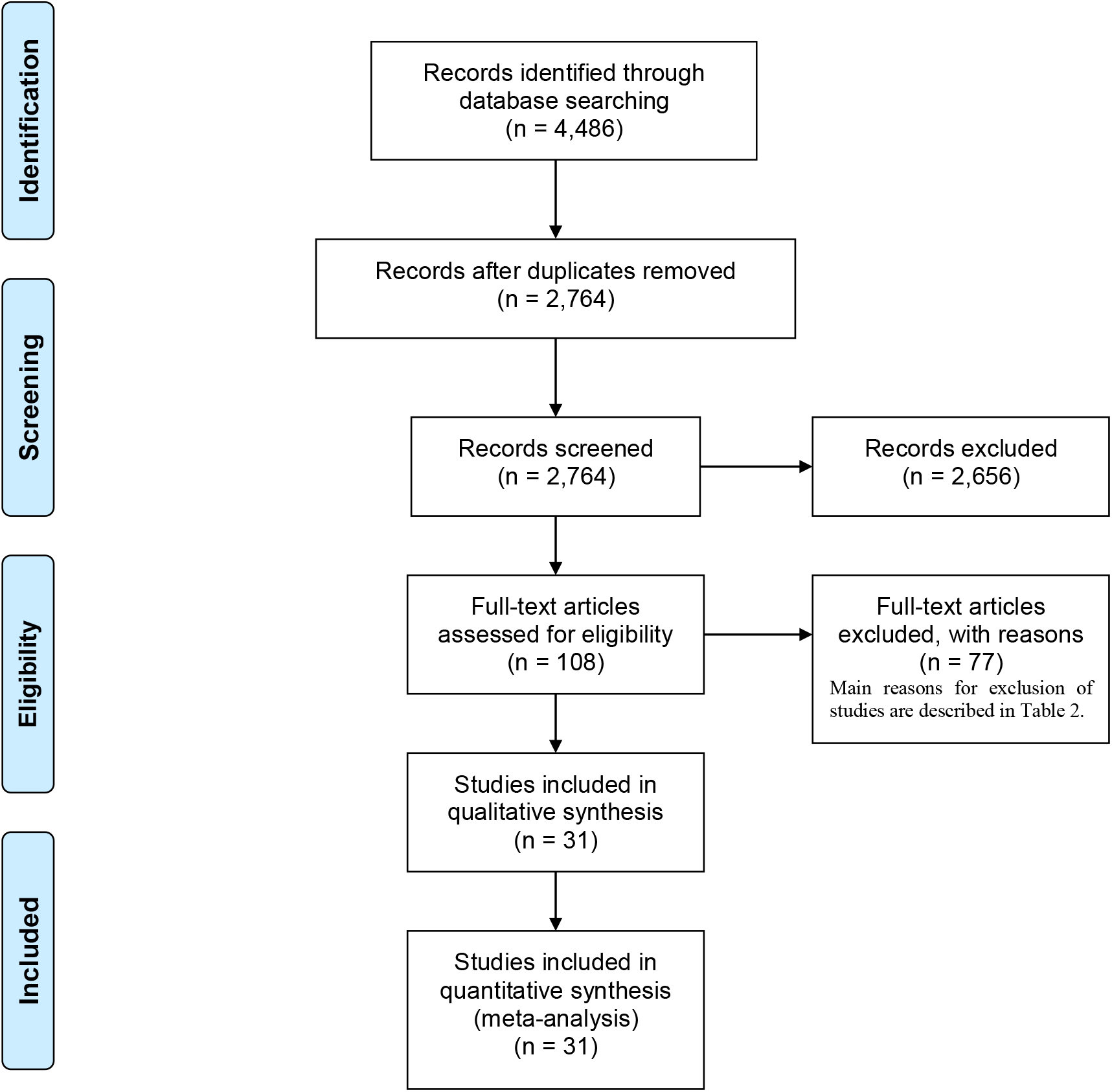

A total of 72,577 individuals 18 years of age or older composed the sample of this systematic review. Twelve studies were performed only with younger adults, while sixteen studies comprised older adults as part of the adult sample. Only three studies were performed only with older adults^18,28,32^, and one including participants aged 65 and over^18^.

The studies included in the meta-analysis were published between 1991 and 2020. Most studies were carried out in high-income countries. European populations represented most of the targeted participants (18 out of 31 studies). Only one study was conducted in Asia^19^, five studies were conducted in Oceania^20-24^ and seven in North America^18, 25-29^. In relation to country’s economy, the included studies were diverse. There were studies from low-income economies, such as India^19^, and one study had its sample composed of Chinese and Danes participants, middle and high-income countries, respectively^30^.

Considering the 31 manuscripts, 24 studies had used validated scales of dental fear and anxiety (DFA) and seven used a single question as DFA assessment tool. When the DFA nomenclature as reported by the article was analyzed, fifteen studies referred to dental anxiety^18, 19, 24, 25, 27, 28, 30-37^, eleven referred to dental fear^9, 10, 21-23, 26, 38-41^, three referred to dental fear and anxiety^20, 29, 42^, while four referred to dental phobia^43-46^. In relation to methods of DFA evaluation, the following tools were used: Dental Anxiety Scale (DAS), Modified-Dental Anxiety Scale (MDAS), Dental Anxiety Question (DAQ), Dental Fear Scale (DFS), Gatchel Fear Scale (Gatchel FS), Seattle Item Question, Single-item dental anxiety and fear (SIDAF) and single-item DSM IV. In relation to the quality assessment of the included studies, only three studies presented low risk of bias^44-46^, with the remaining 28 studies presenting high risk of bias.

The global estimated prevalences of any DFA, high DFA and severe DFA in adults were 13.8%, 11.2% and 2.6%, respectively (Figures 2, 3 and 4, respectively). Sensitivity analysis demonstrated that the exclusion of any one study would not significantly modify the estimated prevalence of each DFA level assessed in this systematic review (data not shown).

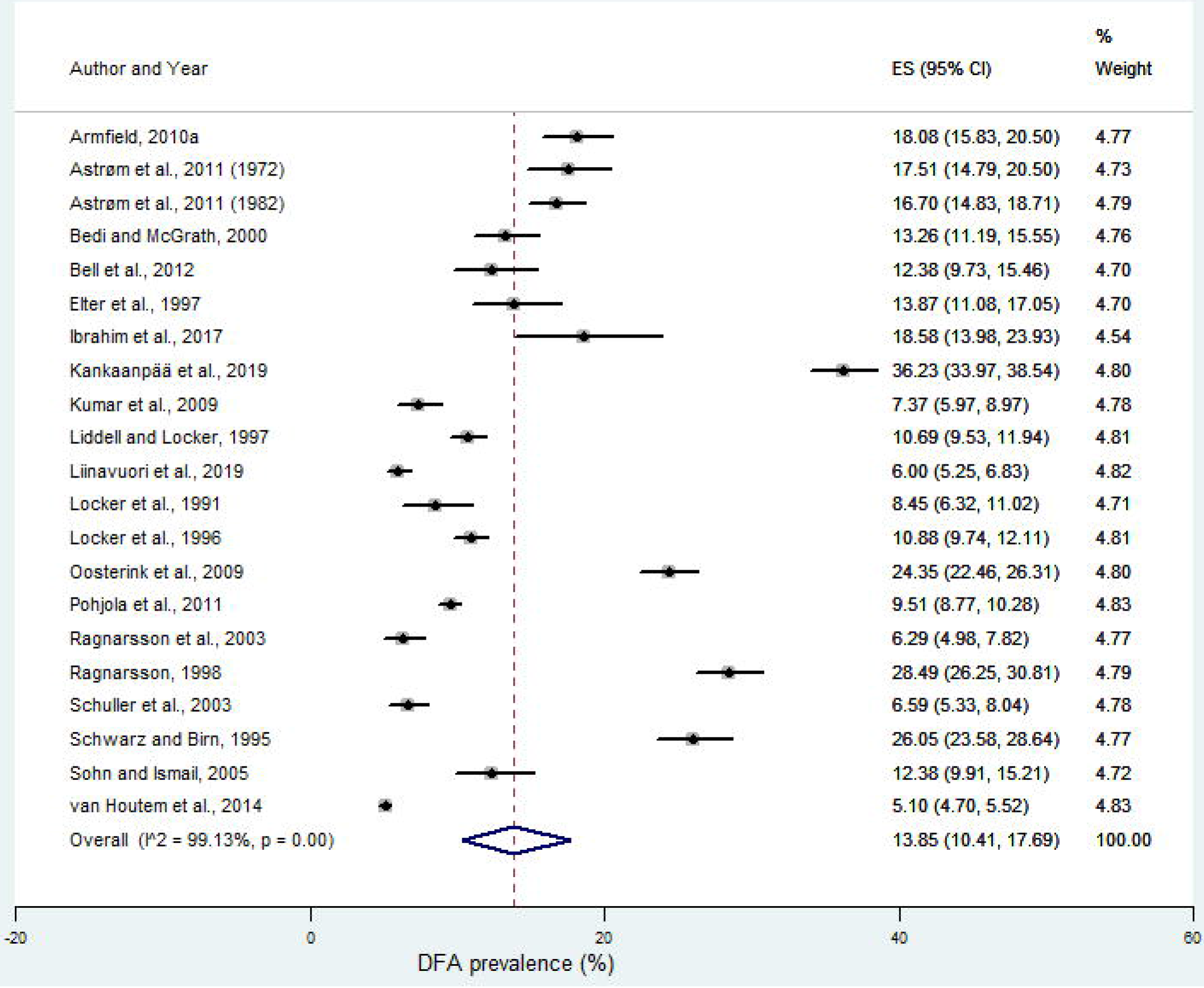

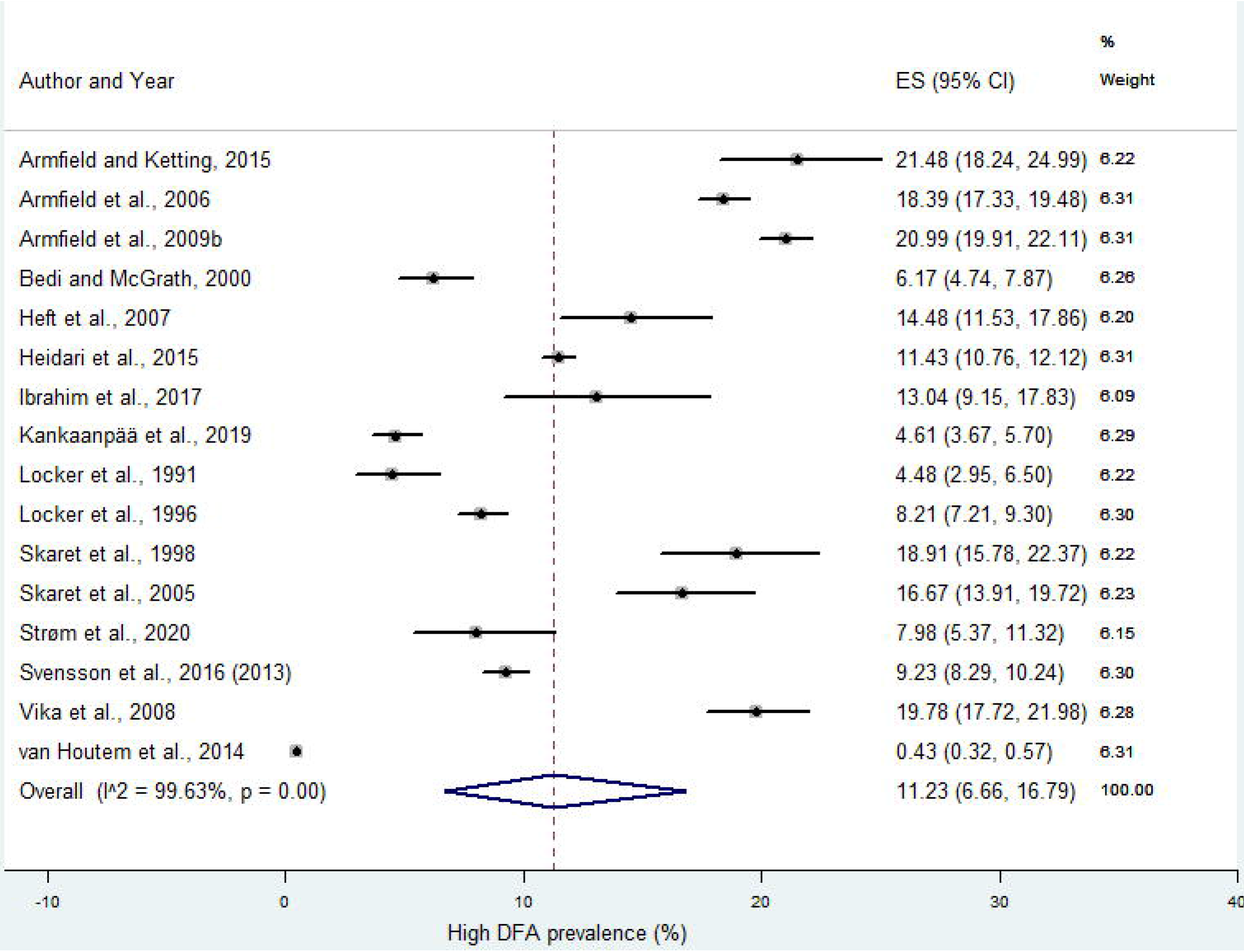

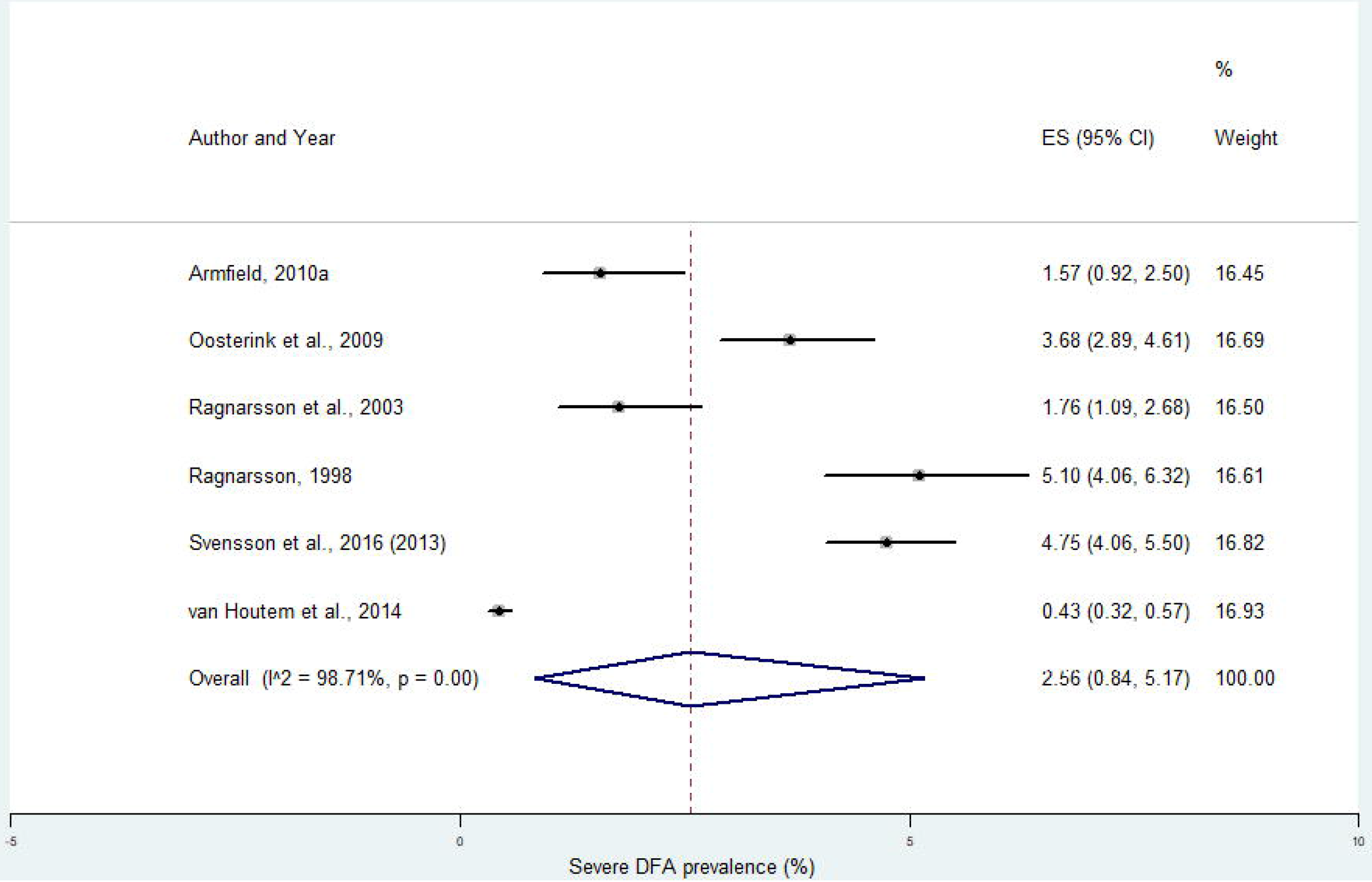

Subgroup analysis of dental fear by gender showed a higher prevalence among women (18.3%) (Supplementary Figure 2) while men reported a prevalence of 10% (Supplementary Figure 3). In relation to high DFA (Supplementary Figure 4) and severe DFA (Supplementary Figure 5), the estimated prevalence for women were 18.6% and 2.8%, respectively. For men, estimated prevalence of high and severe DFA of 9.2% and 2.5% were observed (Supplementary Figures 6 and 7, respectively).

When comparing tools for DFA assessment, the method employed showed high variability. The most used instruments were the DAS and DAQ. Estimated prevalence of 12.7% among studies using DAS and 13.4% among those using DAQ were found. When High DFA was considered, estimated prevalence of 5.5%, 14.3% and 15.7% were observed for the DAS, the DAQ and the DFS, respectively. In relation to severe DFA, when extreme categories are considered, for the DAS and the DAQ a prevalence of 1.6% and of 4.8% were found (Supplementary Figures 8, 9 and 10, respectively).

In relation to the estimated prevalence of DFA according to age, subgroup analysis did not show significant differences (p=0.943). Younger and older, separately, both showed a DFA prevalence of almost 14%. When adults and older adults were evaluated together, an estimated prevalence of 13.2%. was observed (Supplementary Figure 11). High DFA prevalence of 12.3%, 11% and 6.2% (were found for younger adults, older adults, and both in the same age group, respectively (Supplementary Figure 12). Prevalence of severe DFA was estimated to be 3.8% among adults and 2.3% among older adults (Supplementary Figure 13).

## DISCUSSION

The global prevalence of dental fear in this review was high for both any DFA (13.8%) and high DFA (11.2%). Severe DFA reached the percentage of 2.6%. Considering the negative effect of the vicious cycle of dental fear to oral health^1, 2^, the high prevalence of dental diseases^13^ and their significant financial impact for both individuals and health systems^47^, these figures are concerning. These findings reinforce the need to address individual and collective strategies to prevent or treat dental fear.

This review investigated dental fear among adults of varying ages, including older adults separately. Dental fear usually arises in childhood, often related to negative experiences, and it can persist during the life course^48^. Indeed, evaluating dental fear across the lifecourse, into adulthood, Thomson et al.^3^ observed six different trajectories of dental fear, with the onset of dental fear happening even in adulthood. In this review, most of the original studies had a cross-sectional design and therefore did not investigate the occurrence of dental fear in previous stages of life or its onset. When considering the age of individuals, we assessed studies evaluating younger adults and older adults together, only older adults and only younger adults. The prevalence of dental fear was higher in studies that considered both adults and older adults, instead of those only evaluating one age group. Most studies included both younger and older adults, without reporting the number of individuals in each group. This limited our ability to evaluate a trend of dental fear with age and this was a limitation of this review. Also, the number of studies including only adults or elders was small and most studies presented a high risk of bias. On the other hand, this review comprised comprehensive searches without language restrictions on five electronic databases and grey literature. Also, all steps of the systematic review were performed in duplicate, to minimize individual bias.

Considering severe dental fear, this systematic review demonstrated that, when evaluating only older adults, a lower prevalence of dental fear compared to younger adults was observed. Anxiety facing dental procedures tend to decrease with age and it is suggested that this effect may be due to elderly themselves underestimating their need for dental treatment or the ageing processing itself^29^. It has also been suggested that the decrease of dental fear with age may occur because older subjects had more time for good experiences to neutralize aversive ones^31^. In addition, presence of teeth in elderly can be an important factor influencing dental fear. It has been demonstrated that dental fear decreases with age, even when only edentulous people are considered, prevalence of dental fear remained high^21,49^. In DFA and severe DFA, there was no significant differences between younger and older adults. However, it is important to considerer that there may be considerable differences within younger adults, and even within older adults, which this systematic review was not able to identify. The comparison between younger and older adults here did not look at such things, but the effects may still be operating. Armfield et al. showed important differences related to socioeconomic and demographic characteristics, and self-reported oral health status between individuals with high and low fear^21^. In other hand, Liddell and Locker also raised the point about being edentulous when evaluating age differences in dental fear. In their sample, even though older adults were more often edentulous than younger ones, the majority had some teeth remaining. The authors suggested that dental fear in older individuals had not diminished^21, 31^.

In the present study a higher prevalence of any dental fear and high dental fear was observed in women. This is in accordance with the scientific literature. One possible explanation for this finding is based on cultural patterns and social desirability. Some people may see consider it to be more culturally acceptable for women to express fear, while for men the cultural pattern in most societies is that they should not express pain or fear^50,51^. Indeed, in pre-school children, at which time social behaviors are still not set, girls and boys tend to express similar levels of dental fear^50^.

The original studies included in this review used a range of instruments to measure dental fear/anxiety. While it is positive that researchers have different options to investigate dental fear, the use of different scales and measures and different cut-offs generates high heterogeneity between studies, making difficult to compare results. Locker et al^25^ used three different instruments to investigate fear in Canadian adults. While Corah’s DAS presented a prevalence of 10.9%, the Milgrom single item showed 21.0% and the Gatchel’s ten-point fear scale 8.2%, exhibiting a great variation on dental fear prevalence according to the chosen instrument. As a solution, Schuurs et al (1993)^52,53^ suggested that the most reliable results when investigating dental fear are achieved when using more than one questionnaire, including a non-anxiety questionnaire as well. The use of a validated single question to determine dental fear/anxiety (DAQ) has been largely used in epidemiological surveys, especially considering that we only included population-based studies. Usually, epidemiological studies tend to investigate a series of outcomes and the use of shorter and validated instruments are preferred to reduce the time of interview/examination. More complex scales or questionnaires are more difficult to use in large epidemiological studies investigating different oral health outcomes, being more feasible when investigating only a few outcomes or on a smaller sample.

The findings from the present study could be inferred for the world population with caution. Most of the original studies included in the review came from developed countries (mainly based in Europe, North America, and Oceania), with few investigations coming from developing countries. Socioeconomic and cultural aspects may differ from developed and developing countries and these aspects have been demonstrated to influence oral health outcomes.

In this review we considered different terminologies, dental fear, dental anxiety, or dental fear and anxiety, to be the similar. It is important to highlight that, conceptually, they refer to different states of emotions. Dental fear is a feeling provoked by a real, present, specific stimulus (eg, needles or drills), while in anxiety the threat is unclear or not immediately present. Still, the emotional responses of individuals are virtually the same in both situations, and the terms anxiety and fear are often used as synonyms in the literature.

## CONCLUSION

Dental fear and high dental fear are highly prevalent in adults worldwide, being more prevalent among women. Since dental fear is a barrier for regular dental treatment and could lead to worsening of oral health conditions, interventions to prevent and treat dental fear should be provided by clinicians and dental health services, which would improve people’s psychological and oral health.

## Data Availability

All data referred to in the manuscript are availability with the authors.

## ACKNOWLEDGEMENTS

ER Silveira received a pos-doctoral scholarship (PDJ) from CNPq. HS Schuch holds a scholarship (Young Talent) from UFPel Capes Print (Institutional Program for Internationalization). MG Cademartori received a pos-doctoral scholarship (PNPD) from Capes. FF Demarco are supported by fellowships from the National Council for Scientific and Technological Development (CNPq), Brazil.

