## Supplementary Figure Table 1 for "Estimated prevalence of dental fear in adults: a systematic review and meta-analysis"

**Supplementary Table 1.** Excluded studies and main reasons for exclusion. 2020.

| Study | Setting | Reason for exclusion |
|---|---|---|
| Armfield et al., 2007 | Australia | Individuals under 18 years of age |
| Armfield et al., 2008 | Australia | Same sample of Armfield et al., 2009b |
| Armfield et al., 2009a | Australia | Same sample of Armfield et al., 2009b |
| Armfield et al., 2011 | Finland | Same sample of Liinavuori et al., 2019 |
| Armfield et al., 2014 | Switzerland | Individuals under 18 years of age |
| Armfield, 2010b | Australia | Data missing of dental fear prevalence |
| Armfield, 2011a | Australia | Same sample of Armfield, 2010a |
| Armfield, 2011b | Australia | Same sample of Armfield, 2010a |
| Bernabé et al., 2017 | UK | Individuals under 18 years of age |
| Boman et al., 2012 | Sweden | Specific sample |
| Bommireddy et al., 2016 | India | Non-representative sample |
| Bonafé and Campos, 2016 | Brazil | Data missing of dental fear prevalence |
| Carlsson et al., 2015 | Sweden | Same sample of Svensson et al., 2016 |
| Carvalho et al., 2012 | Brazil | Specific and non-representative sample |
| Costa et al., 2018 | Brazil | Specific sample |
| Dixon et al., 1999 | New Zealand | Data missing of dental fear prevalence |
| Doerr et al., 1998 | USA | Non-representative sample |
| El Rey et al., 2005 | Brazil | Specific sample |
| Enkling et al., 2006 | Germany | Non-representative sample |
| Fiset et al., 1989 | USA | Non-representative sample |
| Forslund et al., 2002 | Sweden | Non-representative sample |
| Furuta et al., 2012 | USA | Specific sample |
| Gatchel et al., 1983 | USA | Non-representative sample |
| Gatchel, 1989 | USA | Non-representative sample |
| Goettems et al., 2014 | Brazil | Specific sample |
| Hägglin et al., 1996 | Sweden | Specific sample |
| Hägglin et al., 1999 | Sweden | Specific sample |
| Hägglin et al., 2000 | Sweden | Specific sample |
| Hakeberg et al., 1992 | Sweden | Non-representative sample |
| Hällström and Hägglin, 1984 | Sweden | Specific sample |
| Halonen et al., 2012 | Finland | Specific sample |
| Heidari et al., 2017 | UK | Same sample of Bernabé et al., 2017 |
| Holde et al., 2016 | Norway | Data missing of dental fear prevalence |
| Hu et al., 2007 | Brazil | Specific sample |
| Humphris et al., 2013 | UK | Same sample of Bernabé et al., 2017 |
| Jonsson et al., 2020 | Norway | Data missing of dental fear prevalence |

| | | |
|---|---|---|
| Kirova et al., 2010 | Bulgaria | Non-representative sample |
| Lahti et al., 2007 | Finland | Same sample of Liinavuori et al., 2019 |
| Liinavuori et al., 2015 | Finland | Same sample of Liinavuori et al., 2019 |
| Locker and Liddell, 1991 | Canada | Non-representative sample |
| Locker and Liddell, 1995 | Canada | Non-representative sample |
| Locker et al., 1997 | Canada | Dental anxiety prevalence considering three measures |
| Locker et al., 1999a | Canada | Same sample of Locker et al., 1997 |
| Locker et al., 1999b | Canada | Same sample of Locker et al., 1997 |
| Locker et al., 2001b | New Zealand | Same sample of Locker et al., 2001a |
| McGrath et al., 2001 | UK | Individuals under 18 years of age |
| McGrath et al., 2004 | UK | Individuals under 18 years of age |
| Mellor, 1992 | UK | Specific sample |
| Meneses et al., 2014 | Brazil | Specific sample |
| Meng et al., 2007 | USA | Same sample of Heft et al., 2007 |
| Milgrom et al., 1988 | USA | Non-representative sample |
| Moore et al., 1988 | Denmark | Individuals under 18 years of age |
| Moore and Brødsgaard, 1995 | Denmark | Individuals under 18 years of age |
| Neverlien, 1990 | Norway | Non-representative sample |
| Ng and Leung, 2008 | China | Specific sample |
| Nicolas et al., 2007 | France | Specific sample |
| Oliveira et al., 2014 | Brazil | Specific sample |
| Pohjola et al., 2007 | Finland | Same sample of Pohjola et al., 2011 |
| Pohjola et al., 2008 | Finland | Same sample of Pohjola et al., 2011 |
| Pohjola et al., 2008b | Finland | Same sample of Pohjola et al., 2011 |
| Pohjola et al., 2009 | Finland | Same sample of Pohjola et al., 2011 |
| Pohjola et al., 2013 | Finland | Same sample of Pohjola et al., 2011 |
| Pohjola et al., 2014 | Finland | Specific sample |
| Pohjola et al., 2016a | Finland | Specific sample |
| Pohjola et al., 2016b | Finland | Specific sample |
| Poulton et al., 1997 | New Zealand | Same sample of Locker et al., 2001a |
| Shahid and Freeman, 2019 | UK | Individuals under 18 years of age |
| Skaret et al., 2003a | Norway | Same sample of Skaret et al., 2005 |
| Skaret et al., 2003b | Norway | Same sample of Skaret et al., 2005 |
| Stouthard and Hoogstraten, 1990 | The Netherlands | Individuals under 18 years of age |
| Suominen et al., 2017 | Finland | Same sample of Pohjola et al., 2011 |
| Taani, 2001 | Jordan | Specific and convenience sample |
| Thomson et al., 2000 | New Zealand | Same sample of Locker et al., 2001a |
| Thomson et al., 2009 | New Zealand | Same sample of Locker et al., 2001a |
| Vassend, 1993 | Norway | Individuals under 18 years of age |
| Velkar et al., 2016 | India | Individuals under 18 years of age |
| Viinikangas et al., 2007 | UK | Sample of public dental clinics. |
